# Older age, lack of vaccination and infection with variants other than Omicron associated with severity of COVID-19 and in-hospital mortality in Pakistan

**DOI:** 10.1101/2023.01.30.23285170

**Authors:** Muhammad Zain Mushtaq, Nosheen Nasir, Syed Faisal Mahmood, Sara Khan, Akbar Kanji, Asghar Nasir, Uzma Bashir Aamir, Zahra Hasan

**Author notes:** Contributed equally (Muhammad Zain Mushtaq, Nosheen Nasir). **Corresponding author** Zahra Hasan, PhD, Professor, Department of Pathology and Laboratory Medicine, The Aga Khan University, Stadium Road, P.O.Box 3500, Karachi 74800, Pakistan.

## Abstract

**Objectives:** We investigated factors associated with COVID-19 disease severity and in-hospital mortality in a low-middle income setting.

**Methods:** Records of 197 adult COVID-19 patients admitted to the Aga Khan University Hospital, Karachi between April 2021 and February 2022 were reviewed. Clinical data including, that of SARS-CoV-2 variants was collected.

**Results:** The median age of the patients was 55 years and 51.8% were males. 48.2 % of patients had non-severe disease, while 52.8% had severe/critical disease. Hypertension (48%) and diabetes mellitus (41.3%) were most common comorbid conditions. Omicron (55.3%), Beta (14.7%), Alpha (13.7%), Delta (12.7%) and Gamma (3.6%) were identified in patients. The risk of severe disease was higher in those aged above 50 years (OR 5.73; 95%CI [2.45-13.7]) and in diabetics (OR 4.24; 95% CI[1.82-9.85]). Full vaccination (OR 0.25; 95%CI [0.11-0.58]) or infection with Omicron variants (OR 0.42; 95% CI[0.23-0.74]) reduced disease severity. Age > 50 (OR 5.07; 95%CI [1.92-13.42]) and presence of myocardial infarction (OR 5.11; 95% CI[1.45-17.93]) was associated with increased mortality, but infection with Omicron (OR 0.22 95% CI 0.10-0.53]) reduced risk.

**Conclusions:** Vaccination was found to protect against severe COVID-19 regardless of the infecting variant and is recommended especially, in those aged over 50 years and with co-morbid conditions.

## Introduction

COVID-19 (Coronavirus Disease 2019) has been diagnosed in greater than 654 million cases with a global death toll of 6.67 million [1]. Global epidemiology of COVID-19 has varied greatly, with the highest burden of deaths to date observed in the USA, Brazil and India [1]. Low middle-income countries such as Pakistan and those in Sub-Saharan and East Africa have been relatively spared and mortality due to COVID-19 has been comparatively lower or under reported [2].

Pakistan has thus far experienced 5 distinct pandemic waves, the first, between March and July 2020, the second betweenOctober 2020 and January 2021, the third, between March and May 2021, the fourth between July and September 2021 and the fifth between December and February 2022 [3]. As of December 30 2022, COVID-19 has been diagnosed in 1.58 million cases in Pakistan with the number of deaths nearing 31,000 [1]. Of the COVID-19 cases in Pakistan, 591,553 (36%) COVID-19 cases were from Sindh province with 40% reported from the city of Karachi [4]. The case fatality rate (CFR) has been 2% with some regional variations. It is of interest to understand the factors associated with COVID-19 severity in the local context. In an earlier study conducted at our center during the first wave of COVID-19; 30% of patients had severe to critical disease at presentation with age and critical disease (identified by septic shock and multiorgan dysfunction) were independently associated with mortality [5].

COVID-19 vaccination was first introduced at the start of 2021 [6] and rolled out in stages based on both access and availability in different populations. As in other parts of the world, this contributed to slowing down the pandemic, controlling both morbidity and mortality from COVID-19 [7].

Variants carrying mutations in the spike glycoprotein have been described by the World Health Organization to be variants of concern (VoCs) and include, Alpha (B.1.1.7), Beta (B.1.351), Gamma (P.1), Delta (B.1.617.2) and Omicron (B.1.1.529) [8]. VoCs exhibit genetic modifications linked to increased transmissibility, more severe disease, reduced efficacy of vaccinations or medical treatment [9]. A study from Canada reported that Alpha, Beta, and Gamma were the main variants causing COVID-19 in 2020 with Alpha being the most prevalent variant [10]. Studies from Brazil and Singapore had reported increased severity of disease in COVID-19 patients with the Gamma variant and Delta variant respectively [11].

G, L and S clade SARS-CoV-2 strains were shown to be present in the first wave of COVID-19 in Pakistan from March until August 2020 [12]. There was a shift in predominance from Alpha to Delta variants between April and July 2021 and then again from Delta to Omicron variants between December and February 2022 [13]. The Omicron variant which emerged in the latter half of 2021, had more than 50 mutations across the genome, resulting in reduced vaccine efficacy with a greater chance of breakthrough infections [14]. Omicron was found to be more transmissible compared with other variants, with a greater chance of re-infections however, with a reduction in hospital associated mortality [15].

Throughout the pandemic, COVID-19 related mortality in Pakistan remained low when compared to the other countries [16]. However, the clinical outcomes of patients with severe COVID-19 related to the infecting variants remain to be elucidated. We, therefore, assessed the risk factors, clinical presentation and outcomes in relation to the VoC in patients with COVID-19 in Karachi, Pakistan.

## Methods

This was a retrospective cohort study of adults with a PCR confirmed COVID-19 patients admitted to the Aga Khan University Hospital (a 700-bedded tertiary care hospital in Karachi, Pakistan), between April 2021 and February 2022.

All COVID-19 cases had a SARS-CoV-2 PCR positive respiratory sample using the SARS-CoV-2 Cobas 6800 Roche assay, at the AKUH Clinical Laboratories, Karachi, Pakistan. The CT (crossing threshold) value of each positive SARS-CoV-2 sample was noted. The cases chosen for review were those for whom SARS-CoV-2 VoC testing had been conducted as part of a World Health Organization supported ongoing genomic surveillance effort [17].

Data was collected on symptoms at presentation, treatment, duration of illness, outcomes and also, COVID-19 vaccination. Laboratory and radiological data were collected for all cases. COVID-19 severity was categorized into non-severe in patients who have oxygen saturation ≥ 94% on room air, severe in those who have an oxygen saturation of < 94% and require supplemental oxygen support and critical in those who have respiratory failure, shock and/or multiorgan dysfunction [18]. WHO ordinal score for the severity of COVID-19 [19], length of hospital stay and in-hospital mortality were also recorded. Individuals who had received both doses of a two-dose vaccine regimen were categorized as ‘Fully vaccinated’, those who had received one dose of a two-dose vaccine regimen were categorized as ‘Partially vaccinated’ and those who had not yet received a single dose of COVID-19 vaccination were classified as ‘Unvaccinated’.

### Identification of VoC

Isolates were screened for VoC; Alpha, Beta, Gamma, Delta and Omicron lineage associated mutations using a PCR-based approach [13].

### Statistical Analysis

Median and interquartile range was reported for continuous variables such as age and length of stay and frequency and percentages were used to describe categorical variables such as gender and mortality. Comparison between categorical variables was determined using Chi-square test or Fischer exact test where appropriate. P-value of < 0.05 was considered significant. Logistic regression analysis was performed to examine the effect of each exposure variable on the severity of illness and in-hospital mortality and adjusted Odds ratios (OR) and their 95% confidence intervals (CI) were estimated. Data was analyzed using STATA version 12.0.

## Results

### Description of COVID-19 cases

We studied the clinical characteristics and disease outcomes of n=197 COVID-19 patients admitted at AKUH between April 2021 and February 2022, to investigate the relationship between disease outcomes and SARS-CoV-2 infecting variants. Three COVID-19 waves were observed in Pakistan during this study period (Supplementary Figure 1A). COVID-19 mortality was observed to be lower in the period between December 2021 and February 2022, as compared with the earlier two waves (Supplementary Figure 1B). Notably, cases in April 2021 were mostly associated with Alpha variants whilst those in July and August 2021 were associated with the Delta wave in Pakistan [20].

The median age of the patients was 55 years (IQR 34-70 years) and 51.8% (n=102) were males, Supplementary Table 1. Hypertension (48%) and diabetes (41.3%) were the most common comorbid conditions. There were 23 pregnant patients (24.2% of females) and most of them were identified on screening before delivery. Amongst the pregnant women, 8 (34.7%) developed obstetric complications including intrauterine fetal demise in two cases.

Upon admission, 48.2 % of patients had non-severe disease, while 51.8% had severe/critical disease. Fifty percent of patients had received systemic corticosteroids. Most patients survived (70.6%) till the time of discharge while 38 (19.3%)patients died and 20 (10.2 %) left against medical advice (Supplementary Table 1).Vaccination data was available for 154 patients. 46.7% of study subjects were unvaccinated and 30 % were vaccinated, of which all but three individuals were fully vaccinated. Complications occurred in 27.4% patients (Supplementary Table 1). The most frequent complication was acute kidney injury in 14.7%, followed by myocardial infarction in 9.1% and pneumothorax/pneumomediastinum in 3.6% patients.

The Omicron variant was found in 55.3% of patients, with Alpha variants in 13.7%, Beta in 14.7% and Delta in 12.7% of cases. As these were in a relatively smaller number compared with Omicron, we combined cases with Alpha, Beta, Delta and Gamma variants into a non-Omicron group for further comparative analysis.

Vaccination data were available for 154 out of 197 patients. 46.7% of patients were unvaccinated and 30 % were fully-vaccinated, and 1.5% were partially-vaccinated.

### Factors associated with severe COVID-19

We investigated the factors associated with severe COVID-19 by stratifying the study group into those with non-severe as compared with severe/critical disease. Age, clinical characteristics, vaccination status and infecting SARS-CoV-2 variants were all compared between the two sub-groups. Most patients who developed severe disease were older than 50 years (p<0.001) and were 12 (p<0.001) or dischared (p<0.001) and increased length of hospital stay (p<0.001). Vaccination (OR 0.45; 95% CI[0.23-0.88]), p<0.001 was associated with non-severe disease. Infection with the Omicron variant was associated with non-severe disease (OR 0.42; 95% CI[0.23-0.74]), p=0.003.

In a multivariate analysis, the independent risk factors for severe/critical disease were found to be age greater than or equal 50 years of (aOR 5.73; 95% CI[2.45-13.7]) and presence of diabetes mellitus (aOR 4.24; 95% CI[1.82-9.85]) whereas being fully vaccinated (aOR 0.25; 95% CI[0.11-0.58]) were found to be protective against severe/critical disease (Table 2).

**Table 1.**
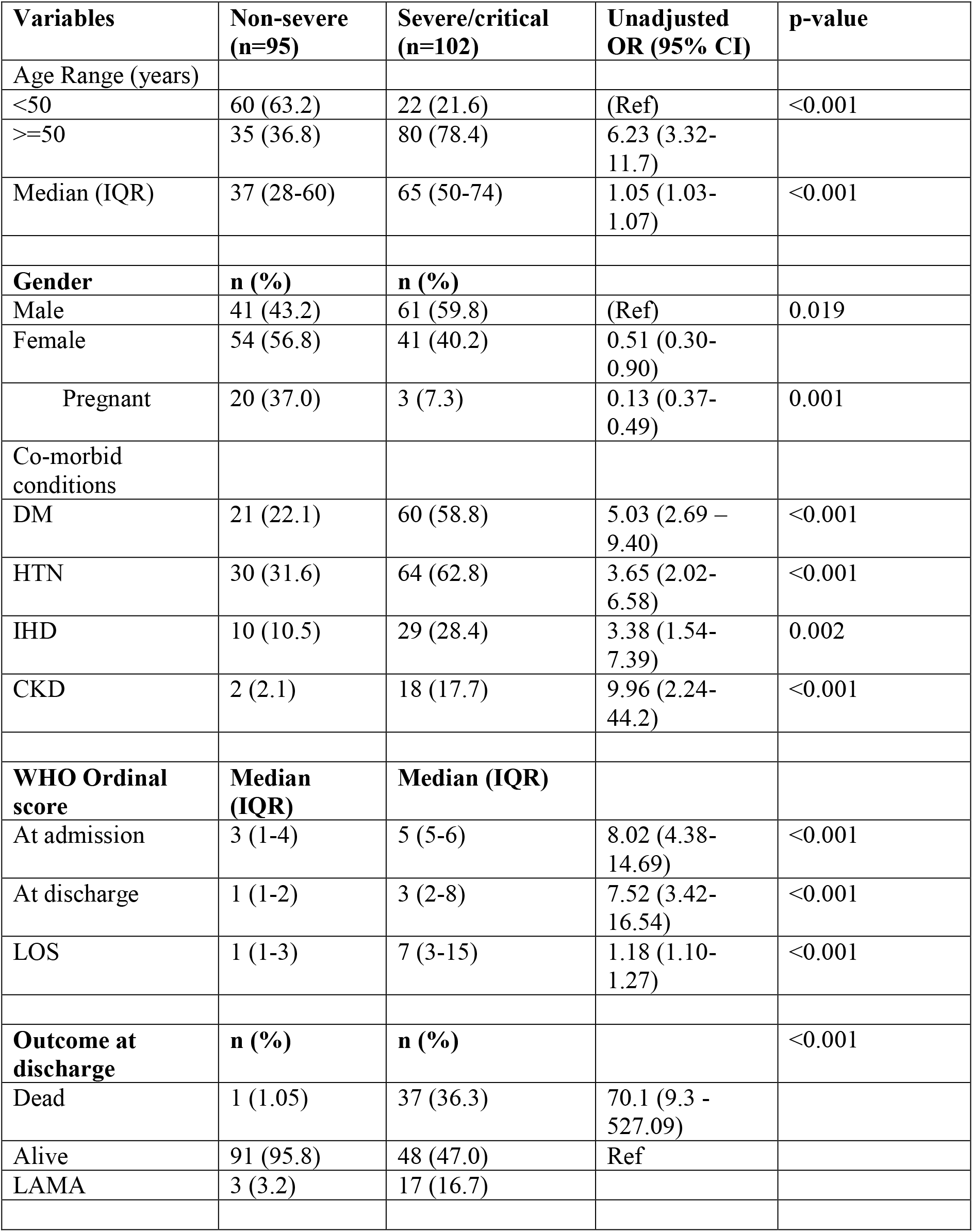
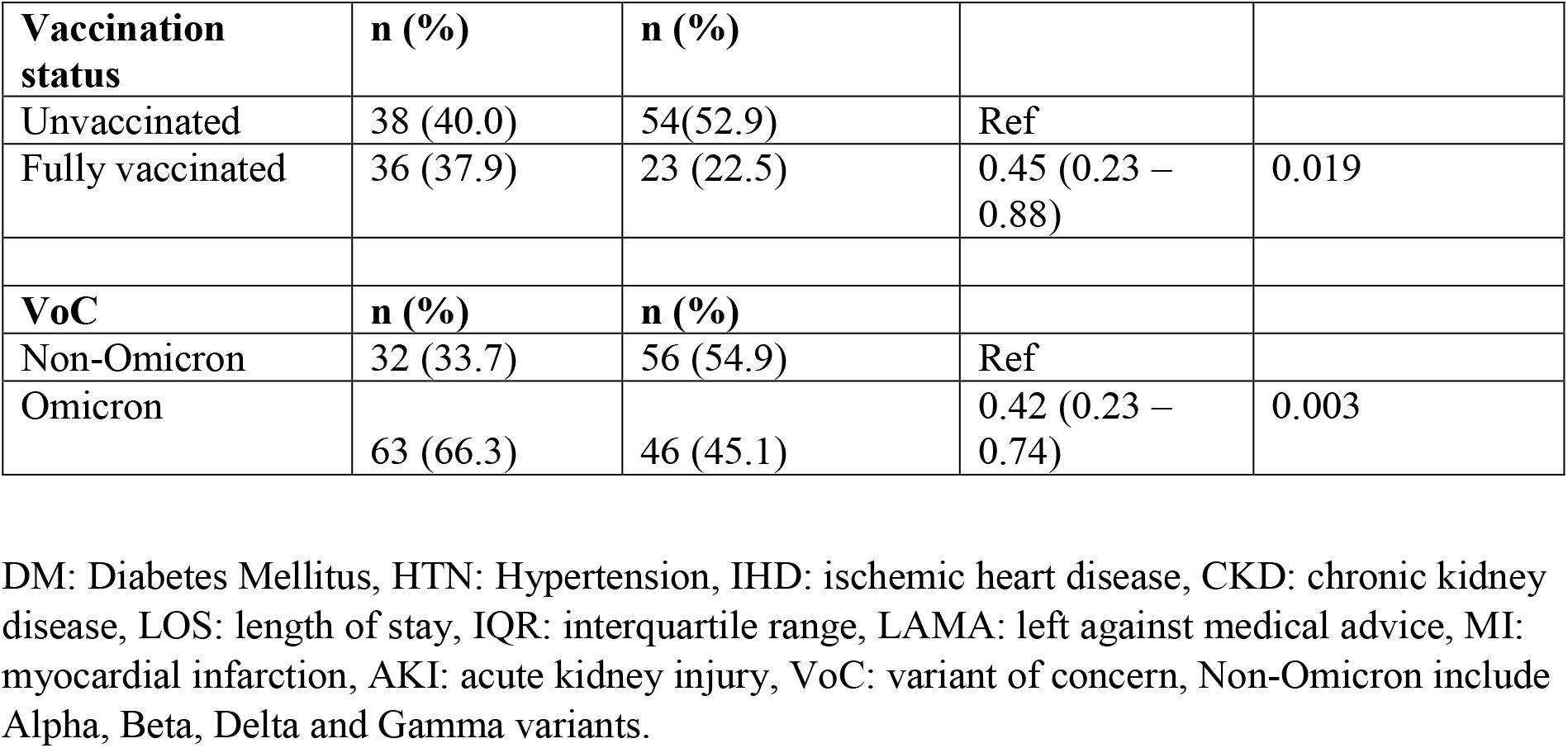
Clinical characteristics associated with non-severe as compared with severe/critical COVID-19.

**Table 2.**
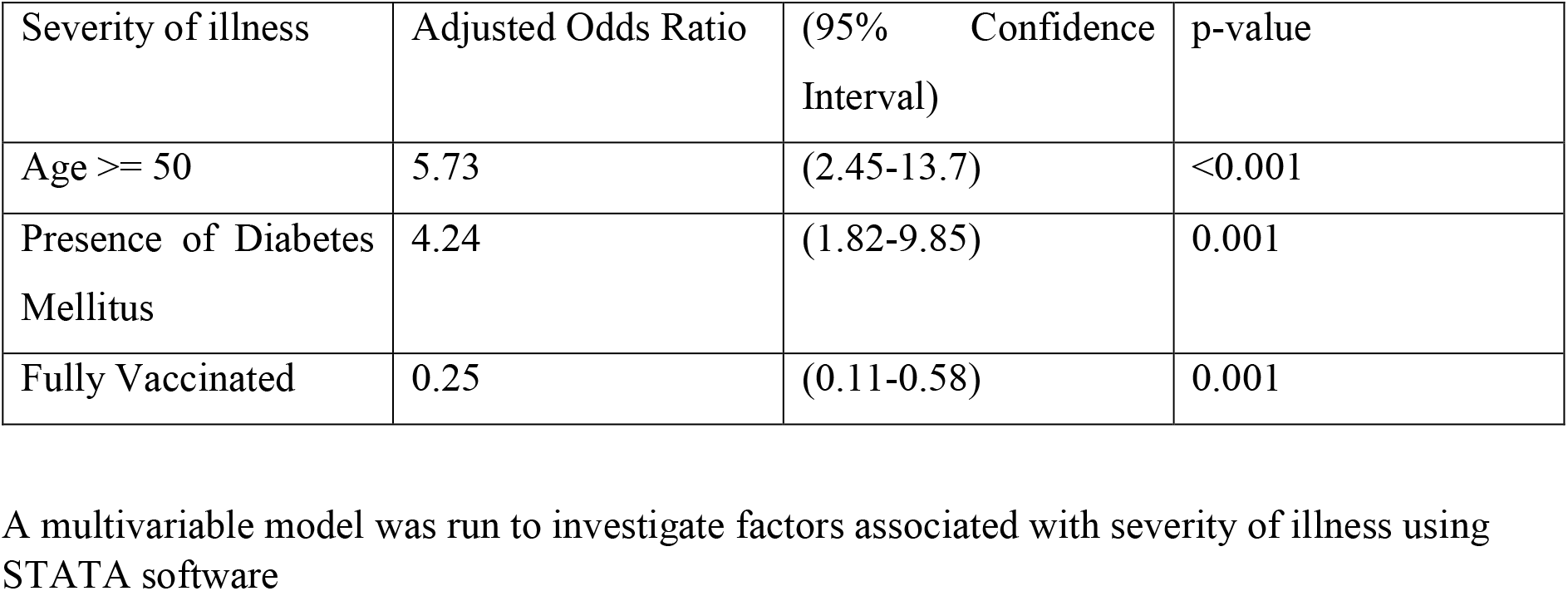
Factors associated with COVID-19 severity.

### Factors associated with mortality

Factors associated with in-hospital mortality in our COVID-19 patient cohort were determined (after excluding 20 patients who left against medical advice, Supplementary Table 1). The median age of those who died was 64 years whilst those who survived was 48 years, Table 3. A univariate analysis showed age greater than or equal to 50 years (OR=5.73; (95% CI: 2.25-14.6), p=0.001; the presence of co-infections or secondary infections (OR:2.45; 95%CI:1.08-5.58), p=0.032, presence of complications such as myocardial infarction (MI) (OR:5.03; 95% CI: 1.69-14.9), p=0.004, acute kidney injury (AKI) (OR:3.78; 95%CI: 1.48-9.61), p=0.005, having pneumothorax or pneumomediastinum (OR:8.05; 95%CI:1.42-45.8), p=0.019 and being unvaccinated (OR: 2.94; 95% CI:1.02-8.39), p=0.044, all to associated with greater risk of death. Infection with Omicron as compared with non-Omicron variants was associated with survival (OR:0.28; 95%CI: 0.13-0.61), p=0.001.

**Table 3:** Clinical characteristics of COVID-19 patients associated with mortality.

| Variables | Died (n=38) | Alive (n=139) | Unadjusted OR (95% CI) | p-value | Multivariate analysis OR (95% CI) | p-value |
| --- | --- | --- | --- | --- | --- | --- |
| **Age Range n (%)** |  |  |  |  |  |  |
| 18-29 | 1 (2.6) | 32 (23.0) | (Ref) |  |  |  |
| 30-49 | 5 (13.2) | 40 (28.8) | 4.0 (0.4-35.9) | 0.219 |  |  |
| 50-69 | 16 (42.1) | 35 (25.2) | 14.6 (1.83-116.6) | 0.011 |  |  |
| >=70 | 16 (42.1) | 32 (23.0) | 16.0 (2.0-127.9) | 0.009 |  |  |
| **Age** |  |  |  |  |  |  |
| <50 | 6 (15.8) | 72 (51.8) | Ref |  |  |  |
| >=50 | 32 (84.2) | 67 (38.2) | 5.73 (2.25-14.6) | <0.001 | 5.07(1.92-13.42) | 0.001 |
| Median Age (IQR) years | 64 (55-75) | 48 (30-69) | 1.04 (1.02-1.06) | 0.001 |  |  |
| **Gender n (%)** |  |  |  |  |  |  |
| Male | 22 (57.9) | 66 (47.5) | (Ref) |  |  |  |
| Female | 16 (42.1) | 73 (52.5) | 0.66 (0.32-1.36) | 0.257 |  |  |
| **Co-morbid n (%)** |  |  |  |  |  |  |
| DM | 20 (52.6) | 49 (35.2) | 2.04 (0.99 - 4.22) | 0.054 |  |  |
| HTN | 22 (57.9) | 59 (42.5) | 1.86 (0.90-3.86) | 0.093 |  |  |
| IHD | 10 (26.3) | 22 (15.8) | 1.90 (0.80-4.46) | 0.141 |  |  |
| CKD | 4 (10.5) | 10 (7.2) | 1.52 (0.45-5.14) | 0.503 |  |  |
| **Co-infection or secondary infections n (%)** | 12 (31.6) | 22 (15.8) | 2.45 (1.08-5.58) | 0.032 |  |  |
| **Complications n (%)** |  |  |  |  |  |  |
| MI | 8 (21.1) | 7 (5.0) | 5.03 (1.69-14.9) | 0.004 | 5.11(1.45-17.93) | 0.001 |
| AKI | 10 (26.3) | 12 (8.6) | 3.78 (1.48-9.61) | 0.005 |  |  |
| Pneumothorax/pneumomed | 4 (10.5) | 2 (1.2) | 8.05 (1.42-45.8) | 0.019 |  |  |
| Laboratory parameters |  |  |  |  |  |  |
| At Day 1 |  |  |  |  |  |  |
| CRP (mg/L)<100 (Ref) | 17 (44.7) | 59 (42.4) | Ref |  |  |  |
| CRP ≥100 | 21 (55.3) | 80 (57.6) | 0.91 (0.44-1.87) | 0.80 |  |  |
| D-Dimer (mg/L)<1.5 (Ref) | 7 (18.4) | 40 (28.8) | Ref |  |  |  |
| D-Dimer >1.5 | 31 (81.6) | 99 (71.2) | 1.78 (0.72-4.39) | 0.204 |  |  |
| **Vaccination status\*** |  |  |  |  |  |  |
| Fully vaccinated | 5 (13.1) | 47 (33.8) | Ref |  |  |  |
| Unvaccinated | 20 (52.6) | 64 (46.0) | 2.94(1.02-8.39) | 0.044 |  |  |
| **VoC n (%)** |  |  |  |  |  |  |
| alpha | 7 (18.4) | 17 (12.2) | (Ref) |  |  |  |
| beta | 8 (21.0) | 18 (12.9) | 1.07 (0.32-0.62) | 0.902 |  |  |
| gamma | 3 (7.8) | 3 (2.2) | 2.42 (0.39 - 15.0) | 0.641 |  |  |
| delta | 8 (21.0) | 15 (10.8) | 1.29 (0.38 - 4.43) | 0.680 |  |  |
| Omicron | 12 (31.6) | 86 (61.9) | 0.34 (0.12 - 0.98) | 0.047 |  |  |
| **VoCs** | **n (%)** | **n (%)** |  |  |  |  |
| Non-Omicron | 26 (68.4) | 53 (38.1) | Ref |  |  |  |
| Omicron | 12 (31.6) | 86 (61.9) | 0.28(0.13-0.61) | 0.001 | 0.22(0.10-0.53) | 0.001 |

DM: Diabetes Mellitus, HTN: Hypertension, IHD: ischemic heart disease, CKD: chronic kidney disease, LOS: length of stay, IQR: interquartile range, MI: myocardial infarction, AKI: acute kidney injury, CRP: C - reactive protein.
\*20 patients who left against medical advice and 3 patients who were partially vaccinated were excluded.

A multivariate analysis run after adjusting for confounding and effect modification from other variables further confirmed in-hospital mortality were found to be associated with age greater than 50 years (aOR 5.07; 95% CI [1.92-13.42]), p=0.001, and the presence of MI (aOR 5.11; 95% CI[1.45-17.93]), p=0.001. Infection with the Omicron variant was associated with survival (aOR: 0.22; 95% CI: 0.10-0.53), p=0.001.

### Viral loads were higher in Omicron infected individuals

Higher SARS-CoV-2 viral loads have been associated with more severe disease. We determined the viral loads present in respiratory specimens and analysed them according to the VoC. The CT of SARS-CoV-2 target gene amplification was used as a marker of viral loads. We found a significant difference between the CT values of the VoC, Figure 1, p=0.015, Kruskal-Wallis analysis. In particular, the CT values of Omicron variants were lower, reflecting significantly higher viral loads as compared with the Alpha variant, p=0.0005.

**Figure 1.**
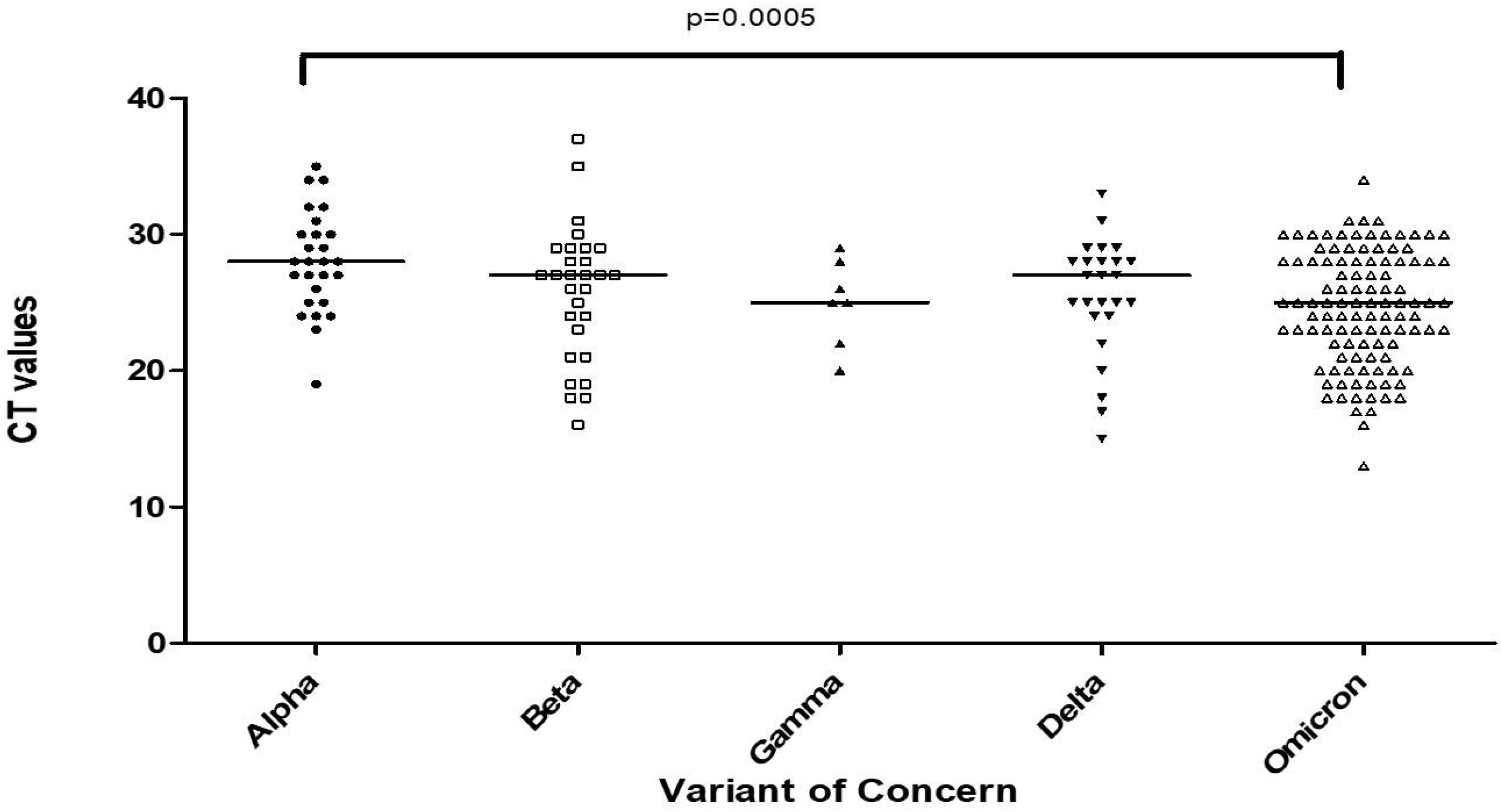
SARS-CoV-2 Omicron variants have lower CT values. The amplification thresholds (CT values) of the infecting VoC in SARS-CoV-2 positive nasal specimens of 197 patients was analysed according to the variant type. The graph depicts data for alpha (n=27), beta (n=29), gamma (n=7), delta (n=25) and omicron (n=109) variants. Data shown is for the Orf1ab gene is depicted as per the SARS-CoV-2 Cobas 6800 Roche assay. The horizontal lines depict the median values for each group. The scatter plots depict interquartile ranges (10-90th percentile) for each group. MWU analysis, with 95% significance p<0.05

## Discussion

We conducted a hospital-based analysis of the association of SARS-CoV-2 VoC with clinical outcomes in patients with COVID-19 from Karachi, Pakistan between April 2021 to February 2022. Diabetes mellitus, hypertension, chronic kidney disease, age over 50 years, and critical disease on presentation were associated with higher mortality. In our cohort, a large proportion of patients were unvaccinated and nearly half developed critical disease. Furthermore, it was noted that infection caused by the omicron variant was associated with significantly lower risk of severe disease at presentation as well as reduced mortality rates.

Omicron variant wave peaked in January 2022 in Pakistan and it was related to highest number of cases since the beginning of the pandemic [1, 21]. In many European countries, the case fatality rate from infections with the Omicron variant was significantly lower as compared with Delta, despite the higher rate of new infections [22]. The period between April and December 2021, was dominated by the Alpha variant then followed by the emergence of the Beta variant in May and the Delta variant in July 2021 [23]. A population based study from UK reported an increase in deaths due to Alpha variants from 2.5 to 4.1 per 1000 detected cases [24].

During the first year of the pandemic (2020) in Karachi, we had observed that severe to critical disease at presentation, and age more than 60 years and critical disease to be independently associated with mortality[5]. This trend remained consistent in 2021 and 2022 except, that patients infected with Omicron variant had lesser odds of dying of COVID-19 as compared with non-Omicron variants, matching previous reports [25]. This fits with reports from South Africa that showed the Omicron wave had a reduced COVID-19 related mortality in patients aged 50 years and above [26].

The majority of our vaccinated study subjects were infected with the Omicron variant likely due to surge in January 2022, and the higher risk of breakthrough infections seen with this variant. Importantly, the Omicron variant had lower CT values associated with higher viral loads as compared with the Alpha variant. This differs from previous variants where genome-wise analysis of SARS-CoV-2 have demonstrated higher viral loads to be associated with increased disease severity [27].

Recent reports show that Omicron variant caused less severe disease across multiple countries in the context of different kinds of COVID-19 vaccinations [15]. In our study, vaccinated individuals were found to have a reduced risk of mortality from COVID-19. This matches previous reports which have shown effectiveness of COVID-19 vaccines in protection against severe disease [7, 28].

A limitation of our study that it is from a single-center tertiary care facility. Further, our sample size was relatively small and we did not have complete vaccination details for all study subjects. Unfortunately, we were unable to collection information regarding vaccination from all cases. During the study period, the most frequently administered vaccines in Pakistan were, BBIBP-CorV (Sinopharm), SinoVac and CanSinoBio. It has been shown that BBIBP-CorV vaccination was protective against severe COVID-19 in Pakistan during the Delta wave [29]. An efficacy study of different vaccines used in Pakistan has shown that inactivated virus type of vaccinations were moderately effective against symptomatic COVID-19, less effective than mRNA vaccination [30]. We were unable to obtain information on the vaccination type from all subjects therefore, our study cannot provide insights on the impact of any specific COVID-19 vaccine. However, this provides key information as to the impact of VoC and its impact on severity of illness and mortality in hospitalized patients with COVID-19 in Pakistan.

In summary, we describe the pattern of SARS-CoV-2 VoCs present in the hospitalized population of COVID-19 cases over the second, third and fourth waves in Pakistan, from April 2021 until February 2022. We observed reduced disease severity and mortality due to the Omicron variant as compared with non-Omicron VoCs. Our findings are in accordance with the trends seen in national data for morbidity and mortality due to COVID-19. Finally, our data reveals that the introduction of VoCs was associated with successive COVID-19 waves. However, presence of Omicron was associated with reduced likelihood of death in our patient population.

## Data Availability

All data produced in the present study are available upon reasonable request to the authors

## Ethics Statement

This work complies with the policies for Ethical Consent as per Helsinki Declaration. The work received approval from the Ethical Review Committee of the Aga Khan University (study reference no. 2021-6232-19404).

## Acknowledgements

Funding for SARS-CoV-2 variant testing was supported by the World Health Organization, Pakistan and through a COVID-19 Research Rapid Grant from the Higher Education Commission, Pakistan.

## Authorship Contributions

The study was designed and supervised by ZH. MZM and NN contributed equally in data analysis and preparation of the manuscript draft. SK, SFM, MZM and NN collected and analysed data. AK and AN conducted the laboratory testing and data analysis. ZH and UBA secured funding for the study. All authors have approved the manuscript submission.

## Contributions

Thanks to Azra Samreen and Ali Raza Bukhari for their technical assistance. Thanks to M Asif Syed and Mansoor Wassan, Department of Health, Sindh, for facilitation with COVID-19 vaccination data.

