## Supplemental Table for "Older age, lack of vaccination and infection with variants other than Omicron associated with severity of COVID-19 and in-hospital mortality in Pakistan"

**Supplementary Table 1. Clinical characteristics of COVID-19 patients**

| **Variable** | **Result** |
| --- | --- |
| **Age Range (years)** | **n (%)** |
| 18-29 | 35 (17.8) |
| 30-49 | 47 (23.9) |
| 50-69 | 59 (30.0) |
| >=70 | 56 (28.4) |
| Median Age (IQR) | 55 (34-70) |
| **Gender** | **n (%)** |
| Male | 102 (51.8) |
| Female | 95 (48.2) |
| Pregnant | 23 (24.2% of females) |
| **Co-morbid conditions** | **n (%)** |
| DM | 81 (41.3) |
| HTN | 94 (48.0) |
| IHD | 39 (19.9) |
| CKD | 20 (10.3) |
| **WHO ordinal score** | **Median (IQR)** |
| At admission | 4(3-6) |
| At discharge | 2(1-5) |
| **Severity of illness** | **n (%)** |
| Non-severe | 95 (48.2) |
| Severe | 39 (19.8) |
| Critical | 63 (32.0) |
| **Medications given** | **n (%)** |
| Systemic steroids | 98 (50.0) |
| Remdesivir | 81 (41.5) |
| Tocilizumab* | 9 (6.1) |
| **Laboratory investigations at admission** | **Median (IQR)** |
| C-Reactive Protein mg/L | 67.9 (23.0-151.1) |
| LDH IU/L | 395 (278-518) |
| Ferritin ng/ml | 349.2 (182-1169) |
| D-Dimer | 1.6 (0.9-4.2) |
| **LOS in days** | 3 (1-9) |
| **Outcome at discharge** | **n (%)** |
| Dead | 38 (19.3) |
| Alive | 139 (70.6) |
| LAMA | 20 (10.2) |
| **Co-infection or secondary infections** | 40 (20.3%) |
| **Complications** | **n (%)** |
| AKI | 29 (14.7) |
| MI | 18 (9.1) |
| Pneumothorax/Pneumomediastinum | 7 (3.6) |
| **Vaccination status** | **n (%)** |
| Fully vaccinated | 59 (30.0) |
| Partially vaccinated | 3 (1.5) |
| Unvaccinated | 92 (46.7) |
| Not known | 43 (21.8) |
| **Cases with VoC** | **n (%)** |
| Alpha | 27 (13.7) |
| Beta | 29 (14.7) |
| Delta | 25 (12.7) |
| Gamma | 7 (3.6) |
| Non-Omicron | 88(44.7) |
| Omicron | 109 (55.3) |

Data describes clinical information for 197 study subjects except ‘*’ where data for only 148 cases was available. DM: Diabetes Mellitus, HTN: Hypertension, IHD: ischemic heart disease, CKD: chronic kidney disease, LOS: length of stay, IQR: interquartile range, LAMA: left against medical advice, MI: myocardial infarction, AKI: acute kidney injury, VoC: variants of concern.
