## Supplemental Figures for "Older age, lack of vaccination and infection with variants other than Omicron associated with severity of COVID-19 and in-hospital mortality in Pakistan"

**Supplementary Figure 1. Description of COVID-19 cases and deaths in Pakistan.** The graphs depict data for COVID-19 between 1 April 2021 and 28 February 2022. A, Cases reported and B, Deaths reported.  Source, John Hopkins, Corona Research Center https://coronavirus.jhu.edu/region/pakistan

A

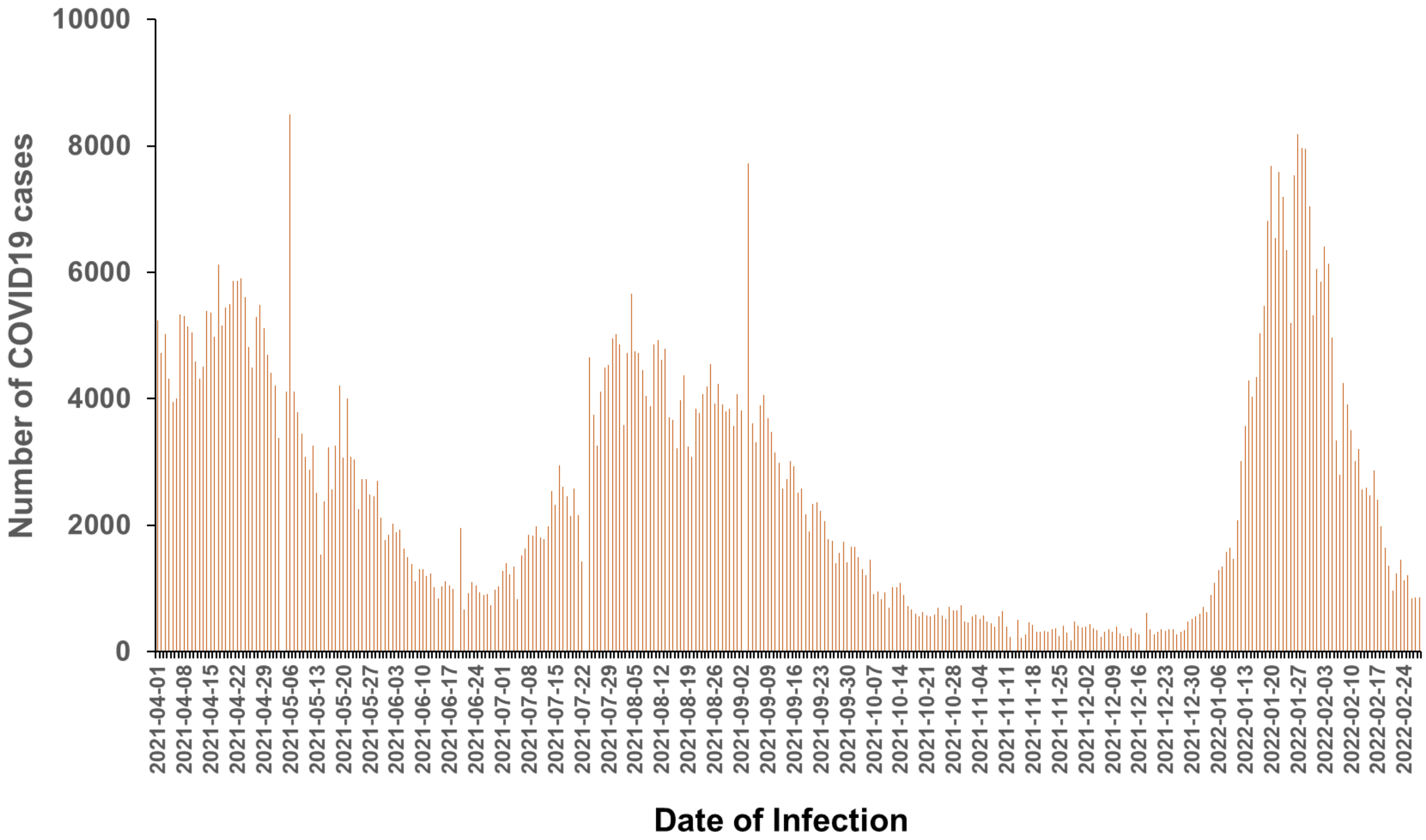

B

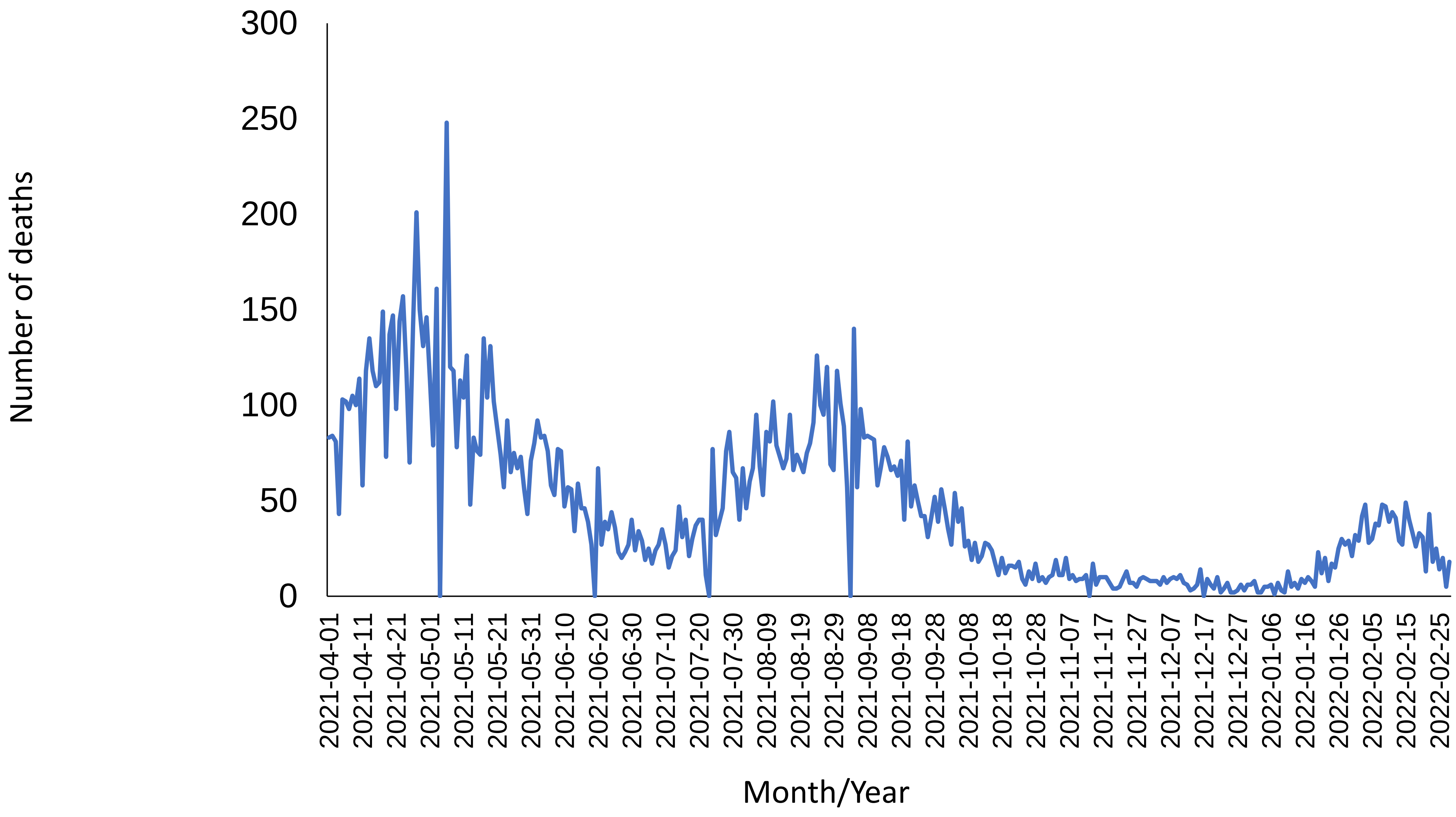

Number of deaths
300
250
200
150
100
50
0
2021-04-01
2021-04-11
2021-04-21
2021-05-01
2021-05-11
2021-05-21
2021-05-31
2021-06-10
2021-06-20
2021-06-30
2021-07-10
2021-07-20
2021-07-30
2021-08-09
2021-08-19
2021-08-29
2021-09-08
2021-09-18
2021-09-28
2021-10-08
2021-10-18
2021-10-28
2021-11-07
2021-11-17
2021-11-27
2021-12-07
2021-12-17
2021-12-27
2022-01-06
2022-01-16
2022-01-26
2022-02-05
2022-02-15
2022-02-25
Month/Year
